# IMPROVING ANTIBIOTICS USE IN PEDIATRIC HOSPITALS IN ARGENTINA: FEASIBILITY STUDY

**DOI:** 10.1101/2024.02.11.24302598

**Authors:** Facundo Jorro-Baron, Cecilia Echave, Viviana Rodriguez, Maria-Jose Aguilar, Romina Balboa, Marina Guglielmino, Florencia Garcia-Causarano, Veronica Del Negro, Patricia Dondoglio, Esteban Falcon, Luz Gibbons, Celeste Guerrero, Ximena Juarez, Analía López, Erika Matteucci, Ana Paula Rodriguez, Emilse Vitar, Javier Roberti, Ezequiel Garcia-Elorrio, Andrea Falaschi

**Author notes:** **Corresponding author:** Facundo Jorro-Baron MD, MsC(c), Researcher, Quality and Safety in Healthcare, Institute for Clinical Effectiveness and Health Policy, 2024 Ravignani, (1414) Buenos Aires Argentina. Pfizer Competitive Grant Program ID 68479861.

## Abstract

**Background:** We aimed to test the feasibility of a multifaceted intervention to enhance the quality of antibiotic prescription by reducing its overuse and increasing the use of narrow-spectrum agents, comprising a range of antimicrobial stewardship strategies in LMIC pediatric hospitals.

**Methods:** We implemented a quality improvement (QI) initiative for the treatment of three groups of infections: acute lower respiratory infections (ALRI), urinary tract infections (UTI), and skin and soft tissue infections (SSTI) in six units of two academic pediatric hospitals. We used an uncontrolled before-and-after design, preceded by a formative phase, to identify barriers and facilitators. The target population was Healthcare workers (HCWs). The strategy comprised an antibiotic audit and feedback, ward- or pathway-specific treatment guidelines, infection-based interventions focused on improving diagnostic accuracy, tailoring therapy to culture results, optimizing treatment duration (antibiotic time out), pharmacy-based interventions, and education.

**Results:** We recruited 617 patients: 249 in the baseline period (BP) and 588 in the implementation period (IP). The patients in the IP group were younger, weighed less, had higher critical care requirements, and had higher ALRI.

With implementation, we observed an increase in antibiotic days of therapy (1051 vs. 831; RR: 1.23 (1.14;1.33); p<0.001). After adjusting for age and place of hospitalization, the differences were significant. This increase was at the expense of a higher use of Access group antibiotics (382 vs. 310; RR: 1.23 (1.14;1.33); p<0.001) and lower use of the Watch group according to the WHO classification (552 vs. 623; RR: 0.89 (0.84; 0.94); p<0.001).

We observed a decrease in antibiotic resistance in the IP group (5% vs. 13%; p<0.001) at the expense of extended-spectrum β-lactamase.

We found no differences in mortality rates between the two periods.

**Conclusion:** Through a QI initiative, the use of antibiotic stewardship programs in pediatric hospitals was shown to be feasible and may improve antibiotic use. We observed a decrease in antibiotic resistance, which may be due to an increase in antibiotic Access group use.

## BACKGROUND

Antimicrobials are the most prescribed medications in pediatrics, with estimates indicating that between 37 and 61% of hospitalized infants and children receive antibiotics^1^. It has been established that 20 and 50% of these prescriptions are potentially unnecessary or inappropriate^2,3^, and many children continue to receive broad-spectrum antibiotics for viral infections or antibiotic courses that are excessively lengthy^4^. The overuse and misuse of antibiotics, inadequate sanitation, low vaccination rates, and insufficient infection prevention and control practices contribute to the high prevalence of drug-resistant infections in low- or middle-income countries (LMICs)^5^.

The misuse of antibiotics has led to antibiotic resistance, posing a serious threat to public health. Infections caused by multidrug-resistant bacteria are associated with higher mortality rates and prolonged hospital stays than those caused by susceptible bacteria^6^. Given the well-documented causal relationship between antibiotic overuse or misuse and the emergence of resistant bacteria, various organizations, including the World Health Organization (WHO), the Infectious Diseases Society of America, and the German Society of Infectious Diseases, have endorsed action plans that emphasize the importance of antibiotic stewardship programs (ASP) to monitor and promote the optimization of antimicrobial use to preserve our antibiotic armamentarium^7,8^. The implementation of ASP pilot program strategies has resulted in estimated annual cost savings of over €330,000^9^. According to recent studies, ASPs have been found to significantly decrease the overall consumption of antibiotics by 19% and the use of restricted antimicrobial agents by 27% within hospitals^10^.

Antimicrobial stewardship programs (ASPs) have been developed to optimize the treatment of infections, reduce infection-related morbidity and mortality, limit the emergence of multidrug-resistant organisms, and reduce unnecessary antimicrobial use^11,12^. Although ASPs have been developed and implemented in some areas of the world, there is a lack of research on LMICs. This may be because most ASPs are still in the development stage and/or not yet widely accepted as standard-of-care strategies^4^ (4). Introducing ASPs in LMICs presents challenges, owing to factors such as limited availability and access to antibiotics, lack of diagnostics, and poor adherence to treatment^13^. Further research is urgently needed to determine the most effective ways to implement ASPs in LMICs, without compromising the quality of care provided to patients^14^.

The rate of antibiotic overuse in a pediatric Argentine population was 35% and was associated with lower respiratory tract, skin, and soft tissue infections^15^. Additionally, there was a problem with the overuse of ceftriaxone, which is classified as a Watch antibiotic in the AWaRe classification system developed by the WHO. In 2019, the prevalence of antibiotic use in one of the included hospitals was 37%^16^. Most of the antibiotic indications were based on empirical evidence, with only 15% driven by microbiological results. Approximately 40% of the days of antibiotic therapy were used to treat hospital-acquired infections (HAI). Compliance with facility guidelines was observed in 57% of cases.

To address performance gaps, quality improvement (QI) initiatives have been employed for several decades to disseminate evidence and to learn from implementation science^17^. Our objective was to assess the feasibility of a multifaceted intervention aimed at enhancing the quality of antibiotic use by reducing overuse and increasing the use of narrow-spectrum agents through implementation of a range of antimicrobial stewardship strategies in pediatric hospitals.

## METHODS

### Study design

We undertook a Quality Improvement (QI) initiative in the inpatient units, neonatal intensive care units (NICU), and pediatric intensive care units (PICU) of two hospitals in Argentina. The hospitals were public, pediatric, and academic, with PICU and NICU level 1 (the best possible level). This project was conducted following a formative phase, following the design of a before-and-after study, encompassing a baseline period (BP) of 22 weeks and an intervention period (IP) of 30 weeks. This initiative was developed by applying the Institute for Healthcare Improvement’s Breakthrough Series Model^18^. QI initiatives are characterized by using healthcare teams to enhance performance on a specific topic through the collection of data and the testing of ideas using plan-do-study-act (PDSA) cycles, supported by coaching and learning sessions. These initiatives are based on the premise that networks of facilities can be transformed into learning systems that accelerate improvements in healthcare performance, with the potential to achieve results on a large scale^19–21^.

### Population

This study targeted healthcare workers (HCWs) from participating hospitals, who were also research subjects. Outcomes were measured in patients admitted to the inpatient units, PICUs, and NICUs of each participating hospital. The study included patients with acute lower respiratory infections (ALRI), urinary tract infections (UTI), and skin and soft tissue infections (SSTI) who were admitted to the participating units.

### Formative Research

Pre-implementation formative research, commonly referred to as formative evaluation, was conducted. This phase employed a qualitative approach. Sixteen semi-structured interviews were conducted with health personnel from the participating units. Employing a rapid qualitative approach, a short-term participatory design was adopted, characterized by the convergence of methods and triangulation of data during data analysis, as well as an iterative process. The interviews were conducted at least one month before the implementation of the intervention and were conducted either in person, by telephone, or online at the convenience of the participants. The interview guide was formulated based on the constructs of normalization process theory (NPT)^22^. The interviews were transcribed and subsequently uploaded to Atlas.ti v8.4, a software program designed for the management of qualitative data.

### Data Management

The Institute for Clinical Effectiveness and Health Policy (IECS) acted as the study data center, providing a specialized data monitoring system to ensure the maintenance of high-quality databases, overseeing all data collection procedures, and facilitating the efficient transfer of study data. A local data center was established at each hospital. The IECS data unit was responsible for monitoring, consolidating, and analyzing the database using Redcap® following Good Clinical Research Practices. The unit also periodically monitored compliance with project procedures, including screening in units, data collection, adherence to the intervention protocol, and data quality. Source data verification was carried out in all participating units, and continuous communication was maintained between site coordinators and data collectors via telephone, email, and WhatsApp®. Run and control charts were shared with teams to monitor progress, and each team reported on the development of improvement opportunities using a standardized report specifically designed for this purpose.

### Outcomes

We evaluated various aspects of antibiotic use and patient outcomes in selected healthcare units. These included the average number of antibiotic days of therapy per 1000 patient days (DOT), proportion of global antibiotic consumption in the Access and Watch groups of the WHO classification^23^, need for antibiotic adjustment, rate of de-escalation (transition from empirical therapy to pathogen-directed therapy based on culture results and clinical guidelines in less than 24 h), adherence to treatment guidelines for infectious diseases, incidence of HAI caused by multidrug-resistant organisms (MDROs), and length of hospital stay in days. We also used in-hospital mortality as a balanced measure, which was defined as a fatal outcome occurring up to hospital discharge.

To adjust for potential confounding factors, we controlled for age, sex, type of infection, and the presence of invasive procedures, such as central venous catheters, urethral catheters, and total parenteral nutrition.

### Intervention

We implemented a complex, implementation science-based package, following a formative research phase. The package was rolled out through two learning sessions and two action periods, utilizing PDSA cycles, and managed by a study coordinator responsible for ASP outcomes. A multidisciplinary team of opinion leaders was assembled for this endeavor^24,25^. Details of the intervention implementation and theory of change are presented in Table 1 and eFigure 1 (Supplement), respectively.

**Table 1.**
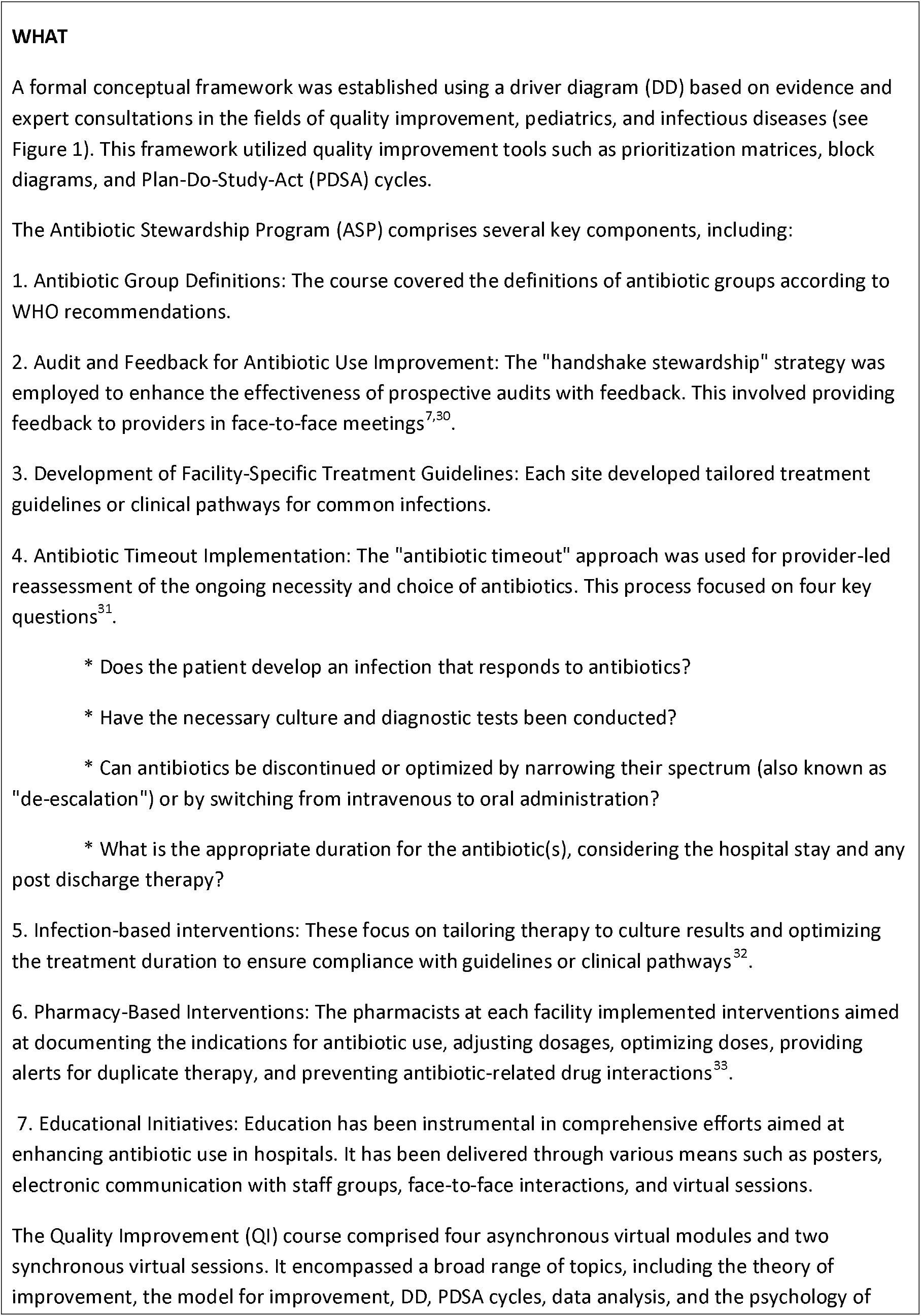
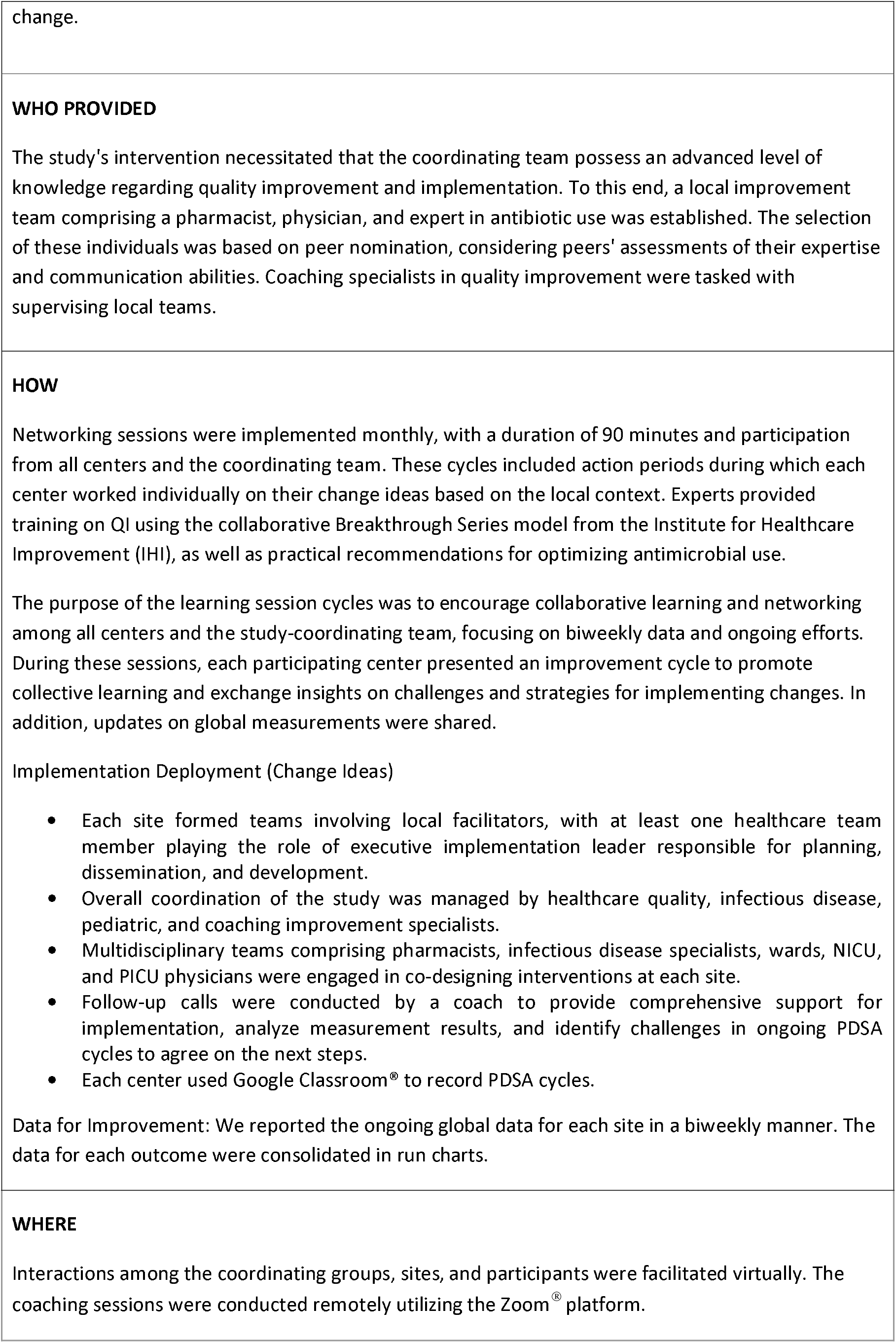
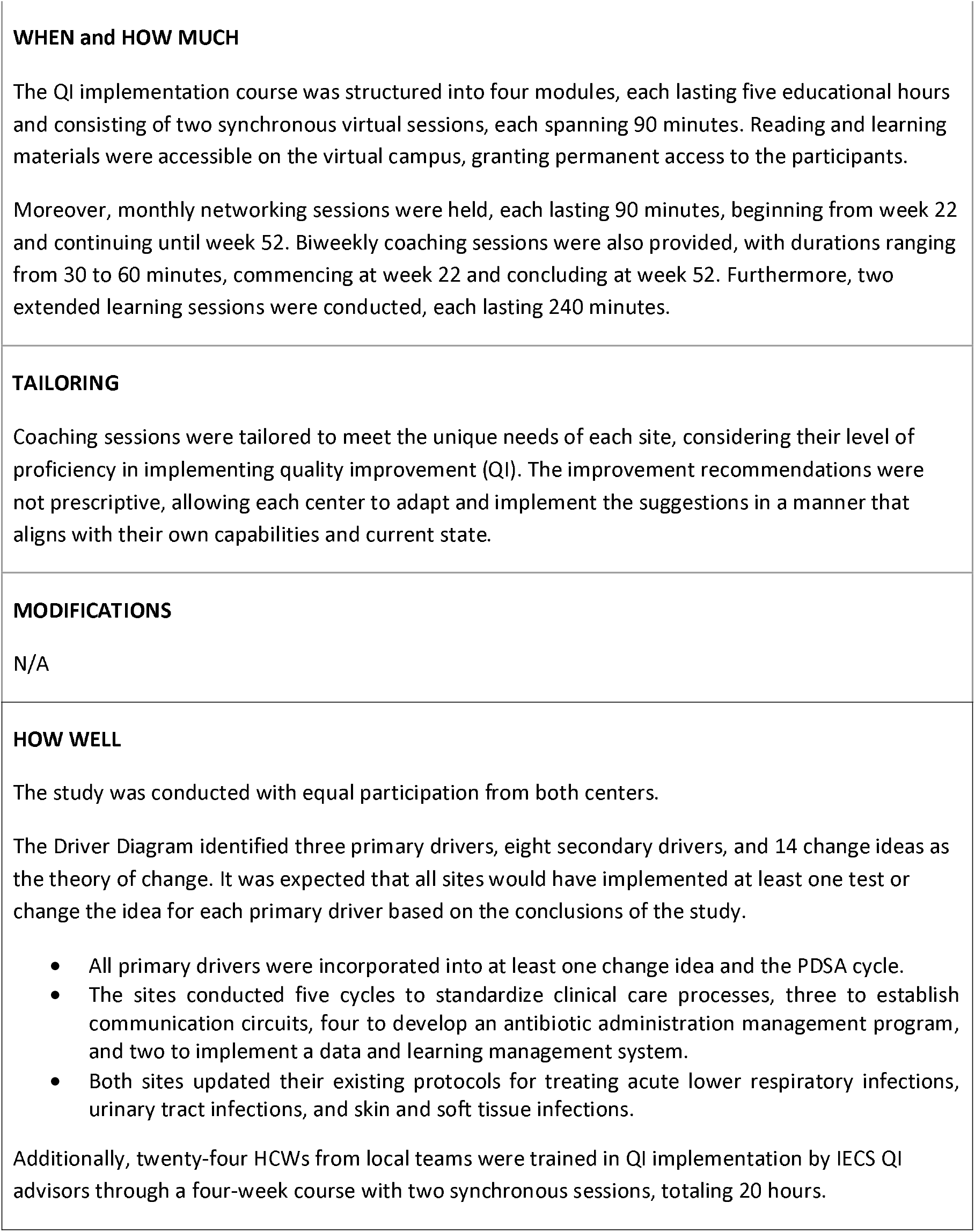
Template for Intervention Description and Replication.

### Statistical Analysis

Patient characteristics were reported during both the study periods. Absolute and relative frequencies were calculated for categorical variables, whereas the median and interquartile range (IQR, quartile 1–quartile 3) were presented for continuous variables. For the comparison between the baseline and implementation periods, the chi-square test or Fisher test was used for categorical variables, and the Wilcoxon rank sum test was used for continuous variables. For the DOT outcome and the number of days of Access and Watch antibiotic consumption, the rate per 1,000 was calculated and reported in the two periods. To test the effect of the intervention, a ratio of the rates was estimated as Rate_IP_/Rate_BP_. To estimate this ratio, a generalized linear model, assuming a Poisson distribution, was used. The same model was used to conduct the subgroup analyses. For the proportion of antibiotic adjustment, de-escalation, and compliance with treatment guidelines, the ratio of the proportions was reported, and binomial distribution was assumed. Data were analyzed using the program R version 4.0.2 (The R Foundation).

### Ethics

The protocol was approved by each hospital IRB. Healthcare workers (HCWs) in the selected units were invited to participate and signed an informed consent request from the local Institutional Review Board (IRB).

## RESULTS

### Formative research

The following recommendations were suggested based on the results of this study: survey the teams responsible for the care of the study patients to enhance the collaborative actions of the intervention. It was recommended that all team members attend training sessions and subsequently participate in the intervention. Potential barriers to intervention, such as differences in criteria and communication issues between the staff, on-call staff, and infectious disease service teams, were identified as significant challenges by all participants. Therefore, it was recommended to restructure the training sessions to ensure that they do not exceed one hour in length and to provide flexible scheduling for asynchronous classes. Additionally, a survey of the technological resources available in each unit should be conducted. Most participants reported connectivity problems and a lack of equipment. It was suggested that specific sessions or meetings be conducted to disseminate the implementation results to enhance the motivation of the participants. In addition, considering the high turnover rate of participants owing to the rotation of residents, it was crucial to implement measures that streamlined the process of training new personnel.

The intervention was adapted according to formative research results.

### Study outcomes

We recruited 249 patients in the BP (1/10/2022-6/5/2022), and 588 in the IP (6/6/2022-1/1/2023). Patients in the IP group were younger (11.5 (3-41) months in the IP group and 30 (9-75) months in the BP group; p<0.001), weighed less (9.1 (5.6-15) kg in the IP group and 12.2 (8-22.3) kg in the BP group; p<0.001), and had higher PICU requirements (46.3% in the IP group and 31.3% in the BP group; p<0.001). Hospital stay was similar between the two groups (8 (5-12) days in the IP group and 8 (4-13) days in the BP group).

The distribution of the type of infection differed between the two groups of patients. The percentages of ALRI in the IP group were 87.0% and 63.2% in the BP group; hospitalization in the PICU was 7.2% and 17.8% in the IP and BP groups, respectively; and the percentages of SSTI were 5.8% and 19.0% in the IP and BP groups, respectively. Patients in the IP group also required more mechanical ventilation (48.6% in the PI and 31.3% in the BPM; p<0.001), more use of central venous catheters (47.6% in the IP and 32.5% in the BP; p<0.001), and higher use of urinary catheters (47.8% in the IP and 24.1% in the BP; p<0.001). The complete characteristics of the patients in both periods are shown in Table 2.

**Table 2.** Characterization of the participating sectors and their patients.

|  | Baseline period<br>(N=249)<br>n/N (%) | Implementation period<br>(N=588)<br>n/N (%) | P-value<sup>§</sup> |
| --- | --- | --- | --- |
| Female sex | 113/249 (45.4) | 254/588 (43.2) | 0.600 |
| Age in months* | 30 (9, 75) | 11.5 (3, 41) | <0.001 |
| Weight (Kg)* | 12.2 (8-22.3) | 9.1 (5.6-15) | <0.001 |
| Unit of hospitalization |  |  | <0.001 |
| Inpatients Unit | 146/249 (58.6) | 265/588 (45.1) |  |
| Pediatric Intensive Care Unit | 78/249 (31.3) | 272/588 (46.3) |  |
| Neonatal Intensive Care Unit | 25/249 (10.0) | 51/588 (8.7) |  |
| Length of stay* | 8 (4, 13) | 8 (5, 12) | 0.700 |
| Discharge place |  |  | <0.001 |
| Home | 163/249 (65.5) | 294/588 (50.0) |  |
| Inpatients Unit | 73/249 (29.3) | 247/588 (42.0) |  |
| Pediatric Intensive Care Unit | 2/249 (0.8) | 27/588 (4.6) |  |
| Other institutions | 4/249 (1.6) | 8/588 (1.4) |  |
| Death | 7/249 (2.8) | 12/588 (2.0) |  |
| Type of infection |  |  |  |
| Acute lower-respiratory infections | 163/258 (63.2) | 556/639 (87.0) | 0.700 |
| Community-Acquired Pneumonia | 123/163 (75.5) | 405/556 (72.8) |  |
| Health Care-Associated Pneumonia | 15/163 (9.2) | 45/556 (8.1) |  |
| Ventilator-Associated Pneumonia | 14/163 (8.6) | 53/556 (9.5) |  |
| Other | 11/163 (6.7) | 53/556 (9.5) |  |
| Urinary Tract Infection (UTI) | 46/258 (17.8) | 46/639 (7.2) | <0.001 |
| Community-Acquired UTI | 26/46 (56.5) | 34/46 (73.9) |  |
| Catheter-Associated UTI | 4/46 (8.7) | 10/46 (21.7) |  |
| Other | 16/46 (34.8) | 2/46 (4.3) |  |
| Skin and soft tissue infections (SSTI) | 49/258 (19.0) | 37/639 (5.8) | 0.999 |
| Community-Acquired SSTI | 41/49 (83.7) | 30/37 (81.1) |  |
| Health Care-Associated SSTI | 7/49 (14.3) | 6/37 (16.2) |  |
| Other | 1/49 (2.0) | 1/37 (2.7) |  |
| Use of Mechanical Ventilatory Assistance | 78/249 (31.3) | 286/588 (48.6) | <0.001 |
| Use of Central Venous Catheter | 81/249 (32.5) | 280/588 (47.6) | <0.001 |
| Use of Urinary Catheter | 60/249 (24.1) | 281/588 (47.8) | <0.001 |
| Use of Parenteral Nutrition | 13/249 (5.2) | 17/588 (2.9) | 0.100 |

\* Median (IQR)
\*\* One patient can have more than one
<sup>§</sup>Wilcoxon rank sum test; Pearson's Chi-squared test; Fisher's exact test

After the implementation of the intervention, there was an increase in antibiotic DOT (1051 in the IP vs. 831 in the BP; RR: 1.23 (1.14;1.33); p<0.001). After adjusting for age and place of hospitalization, this difference remained significant (1.21 (1.16;1.27); p<0.001). The increase in antibiotic DOT was at the expense of a higher use of antibiotics in the Access group (382 in the IP group vs. 310 in the BP group; RR: 1.23 (1.14;1.33); p<0.001), and a lower use of antibiotics in the Watch group (623 in the IP group vs. 552 in the BP group; RR: 0.89 (0.84; 0.94); p<0.001) (Table 3). In the subgroup analysis, we found that age less than one year (RR: 1.56 (1.45;1.68); p<0.001) and more than three years (RR: 1.20 (1.11;1.29); p<0.001), hospitalization in the inpatient unit (RR 1.25 (1.17;1.34); p<0.001), neonatology (RR: 1.38 (1.22;1.57); p<0.001), and compliance with guidelines (RR: 1.38 (1.30;1.46); p<0.001) were significantly associated with increased antibiotic DOT (Table 4). We also found that Access group consumption increased more in the PICU (RR: 1.76 (1.52;2.04); p<0.001) and NICU (RR: 1.43 (1.15;1.79); p<0.001) (Table 3). When we analyzed the ALRI subgroup, we observed more antibiotic DOT in the IP regardless of the unit of hospitalization or age (adjusted RR: 1.08 (1.02;1.14); p=0.006). This increase occurred at the expense of antibiotic consumption in the Access group (adjusted RR: 1.31 (1.19;1.45); p<0.001). In the ALRI group, we also found a decrease in antibiotic consumption in the Watch group during the IP (adjusted RR: 0.85 (0.79;0.91); p<0.001).

**Table 3.**
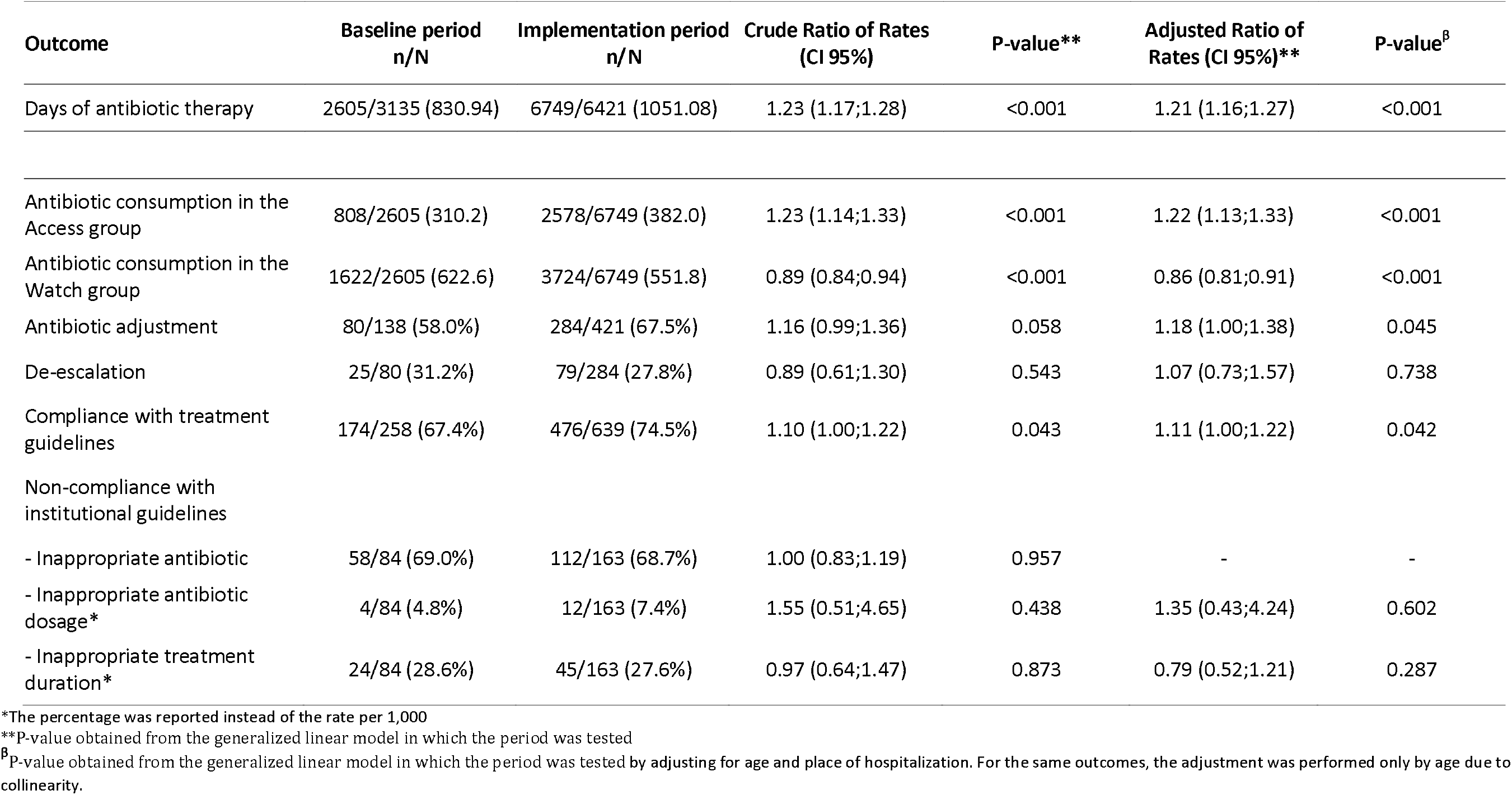
Outcomes by study period and adjustment for age and unit of hospitalization.

**Table 4.** Subgroup analysis of principal outcomes based on hospitalization unit.

|  | Baseline<br>n/N (Rate per 1,000) | Implementation<br>n/N (Rate per 1,000) | Crude Ratio of Rates (CI 95%) | P-value* |
| --- | --- | --- | --- | --- |
| **Days of antibiotic therapy (DOT)** |  |  |  |  |
| **Age Group** |  |  |  |  |
| Less than one year | 846/1417 (597.04) | 3708/3994 (928.39) | 1.56 (1.45;1.68) | <0.001 |
| Between one year and three | 598/592 (1010.14) | 1472/1158 (1271.16) | 1.06 (0.96;1.16) | 0.257 |
| More than three | 1161/1126 (1031.08) | 1569/1269 (1236.41) | 1.20 (1.11;1.29) | <0.001 |
| **Unit of hospitalization** |  |  |  |  |
| Inpatients unit | 1264/1411 (895.82) | 1986/1778 (1116.99) | 1.25 (1.17;1.34) | <0.001 |
| Pediatric intensive care unit | 1004/993 (1011.08) | 3952/3372 (1172.00) | 1.05 (0.98;1.12) | 0.182 |
| Neonatal intensive care unit | 337/731 (461.01) | 811/1271 (638.08) | 1.38 (1.22;1.57) | <0.001 |
| **Compliance with guidelines** |  |  |  |  |
| Yes | 1610/2185 (736.84) | 4774/4698 (1016.18) | 1.38 (1.30;1.46) | <0.001 |
| No | 995/855 (1163.74) | 1962/1679 (1168.55) | 1.00 (0.93;1.08) | 0.916 |
| **Pediatric intensive care + mechanical ventilation** | 921/909 (1013.20) | 3812/3290 (1158.66) | 1.02 (0.95;1.10) | 0.517 |
| **Antibiotic consumption in the Access group** |  |  |  |  |
| **Place of hospitalization** |  |  |  |  |
| Inpatients unit | 504/1264 (398.7) | 825/1986 (415.4) | 1.04 (0.93;1.16) | 0.469 |
| Pediatric intensive care unit | 202/1004 (201.2) | 1401/3952 (354.5) | 1.76 (1.52;2.04) | <0.001 |
| Neonatal intensive care unit | 102/337 (302.7) | 352/811 (434.0) | 1.43 (1.15;1.79) | 0.001 |

\*P-value obtained from the generalized linear model in which the period was tested

Antibiotics that increased their use in the IP were ampicillin (4.3% vs. 7.5%), ampicillin-sulbactam (6.8% vs. 13.8%), gentamicin (1.8% vs. 5.0%), and clarithromycin (15.2% vs. 20.9%), all of which, except for clarithromycin, were antibiotics in the Access group. The new antibiotic guidelines of one hospital incorporated clarithromycin and increased its use locally (19.9% in the BP group vs. 25.1% in the IP group; p =0.050). However, we observed a decreased use of ceftriaxone (16.7% vs. 11.3%), piperacillin-tazobactam (9.9% vs. 6.6%), clindamycin (7.4% vs. 3.0%), and vancomycin (7.8% vs. 5.6%), all of which except for clindamycin were antibiotics in the Watch group (eTable 1, supplement).

We observed that 58% of BP and 67.5% of IP infections required antibiotic adjustments. The de-escalation rate was similar in both periods (31.2% in the BP group vs. 27.8% in the IP group; adjusted RR: 1.07 (0.73;1.57); p=0.738). There was an increase in compliance with the treatment guidelines in the IP group (67.4% vs. 74.5%; adjusted RR: 1.11 (1.00;1.22); p=0.042). The main cause of noncompliance with institutional guidelines was inappropriate antibiotics in both periods (69.0% in BP vs. 68.7% in IP; RR: 1.00 (0.83;1.19; p=0.957) (Table 4).

We could not analyze the rate of HAIs caused by antibiotic-resistant microorganisms because of their low incidence (eTable 2, supplement). However, we observed a decrease in the IP of microorganism antibiotic resistance (13.0% in the BP vs. 5.0% in the IP; p<0.001) at the expense of extended-spectrum β-lactamase microorganisms (61.1% in the BP vs. 47.6% in the IP) (Table 5).

**Table 5.** Microorganism antibiotic resistance by study period.

| Microorganism antibiotic resistance | Baseline period | Implementation period | P-value |
| --- | --- | --- | --- |
|  | (N = 138) | (N = 421) |  |
|  | n/N (%) | n/N (%) |  |
| **Antibiotic resistance** | 18/138 (13.0%) | 21/421 (5.0%) | 0.001 |
| Methicillin resistance | 3/18 (16.7%) | 8/21 (38.1%) | 0.171 |
| Vancomycin resistance | 0/18 (0.0%) | 0/21 (0.0%) | >0.999 |
| Extended-spectrum β-lactamase | 11/18 (61.1%) | 10/21 (47.6%) | 0.523 |
| Carbapenemases | 4/18 (22.2%) | 6/21 (28.6%) | 0.726 |

We found no differences in the mortality rates between the periods (2.8% in the BP group vs. 2.0% in the IP group; p=0.474).

## DISCUSSION

We implemented an ASP program across six units in two pediatric hospitals in a medium-income country utilizing a practical toolkit provided by the WHO (10). Our intervention was based on the guideline recommendation of using >60% antibiotics from the Access group of the AWaRe classification. Although we were unable to achieve this suggested percentage, we observed an improvement in the percentage use of Access group antibiotics, as well as in the subgroup analysis based on the PICU and NICU. This improvement was attributed to enhanced compliance with the observed treatment guidelines.

One of the major strengths of our study was the significant change in the way physicians prescribed antibiotics. We were able to decrease the use of antibiotics within the Watch group and increase the use of antibiotics within the Access group, thereby reducing the development of antibiotic-resistant microorganisms, particularly extended-spectrum β-lactamase microorganisms. In addition, we performed formative research that allowed us to adapt the intervention to each site.

During BP, we evaluated the current level of guideline compliance in each hospital using guidelines that were over 10 years old. We subsequently developed new treatment guidelines for both hospitals, which were strongly supported by local leaders despite the initial resistance from healthcare teams. To standardize clinical care processes and ensure effective implementation of treatment guidelines, hospital teams undertook five PDSA cycles, during which they tested ideas for establishing communication channels, creating an antibiotic administration-management program, and developing a data and learning management system.

Our investigation revealed a significant disparity in DOT between the intervention (IP) and control (BP) groups, which can be ascribed to the increased use of clarithromycin. We found higher rates of clarithromycin prescriptions in one hospital. We recommended this hospital review the use of clarithromycin in the ALRI guidelines. Despite the reformulation of the ALRI guidelines to decrease clarithromycin use, it is important to note that these changes could not be implemented before the completion of the study. A recent systematic review showed that ASPs are not associated with a reduction in antibiotic prescriptions for hospitalized pediatric patients, with only one of the four studies conducted in LMICs^26^. Another report demonstrated little change in antibiotic consumption in LMICs in recent years^27^. Furthermore, a previous analysis of IQVIA data from 2000 to 2010 reported a dramatic increase in global consumption, particularly in BRICS countries (Brazil, Russia, India, China, and South Africa), with this increase continuing through 2015^27,28^. Argentina is currently under evaluation as a member of the BRICS. The need for further research on implementing ASPs in LMICs is crucial because of the numerous obstacles encountered, such as the restricted availability of antibiotics, absence of diagnostic tools, and suboptimal treatment adherence^13,29^. These factors, along with the pressing demand for improved patient care, make it imperative to explore effective methods for implementing ASPs without compromising their quality.

Our findings revealed a decrease in MDRO prevalence in the IP group; however, there were too few cases of multidrug-resistant HAIs to establish a correlation with ASP. We believe that a longer ASP evaluation period is necessary to detect improvements in antibiotic DOT or MDRO incidence.

### Limitations

One of the primary limitations of our study was the discrepancy in the type of infection observed between the two periods. Other limitations included the age and illness severity of the patients, as the BP group consisted of older individuals and the IP group required more PICU admissions. We adjusted the results for age and critical care needs but were unable to improve de-escalation practices. The teams developed PDSA cycles to improve communication between microbiologic results, infectologists, and physicians to change the empirical therapy to pathogen-directed therapy; however, more PDSA cycles may be required to improve. Despite the limited resources available, the data collected during this study were of exceptional quality. They provided highly valuable information regarding the type of antibacterial drug utilized, dosage and duration of usage, culture type and time to rescue, organism resistance patterns, and de-escalation practices.

## CONCLUSION

To the best of our knowledge, this study is one of the first pediatric investigations in South America to evaluate the implementation of ASP in hospitals. Our results suggest that it is practicable to establish an ASP in pediatric hospitals in LMICs, provided that the guidelines set forth by the WHO are followed and a minimum period for assessing the program’s effectiveness can be determined. ASP has the potential to improve the quality of antibiotic use. However, further research is necessary to determine the relationship between ASP and duration of antibiotic use, as longer intervention periods are required for such an assessment.

## Supporting information

https://www.dropbox.com/scl/fi/7g1t0r0hgvilo80hfsu4g/Supplement-SOPA.pdf?rlkey=q14sjamal6388pg6i4ai6anh0&dl=0

## Data Availability

All data produced in the present study are available upon reasonable request to the authors

https://osf.io/dh3xk/?view_only=9799012b018b4db49e94cab6dab0112d

## Acknowledgments

The authors would like to thank the hospitalization staff of the Hospital General de Niños Pedro de Elizalde and Hospital Humberto Notti. We would like to thank Paperpal for the English language editing.

## Data Availability

Link https://osf.io/dh3xk/?view_only=9799012b018b4db49e94cab6dab0112d

