## Supplementary material for "IMPROVING ANTIBIOTICS USE IN PEDIATRIC HOSPITALS IN ARGENTINA: FEASIBILITY STUDY": https://www.dropbox.com/scl/fi/7g1t0r0hgvilo80hfsu4g/Supplement-SOPA.pdf?rlkey=q14sjamal6388pg6i4ai6anh0&dl=0

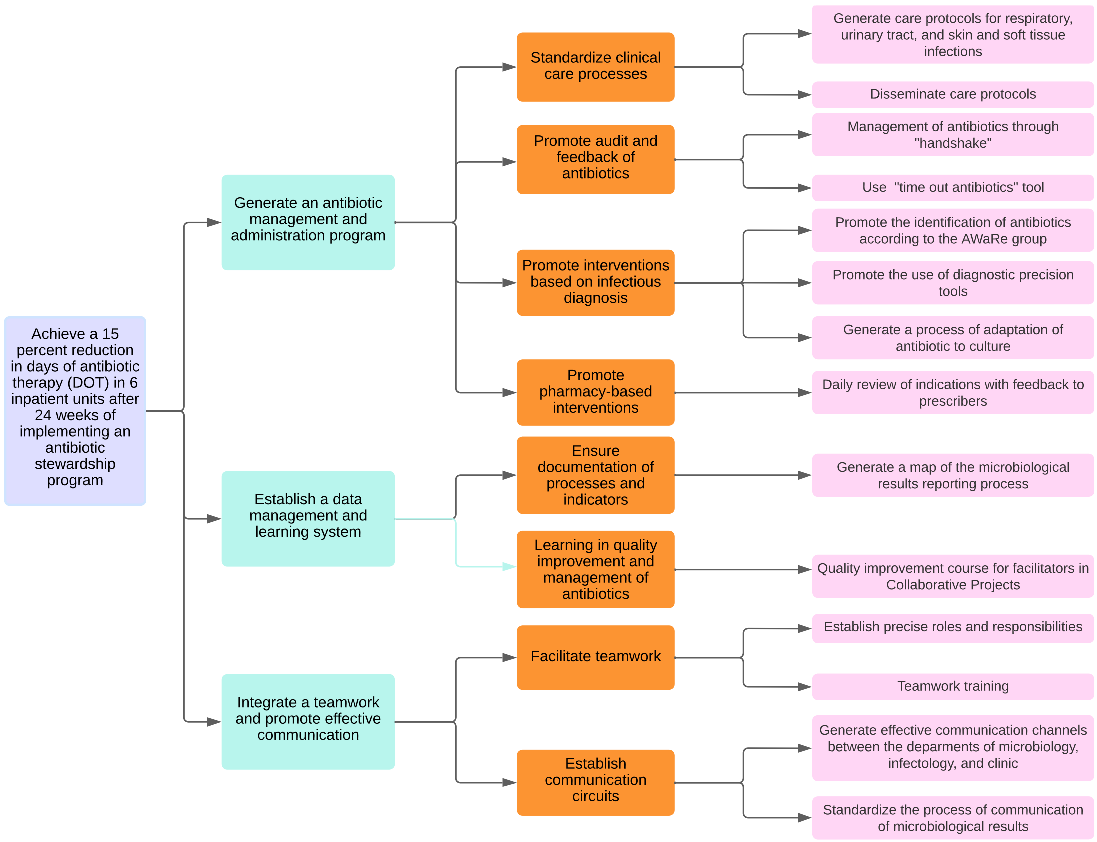

*eFigure 1. Theory of change driver diagram.*

*eTable 1. Antibiotic use and defined daily dose by period.*

| Antibiotic | Baseline period (N= 514) | | Implementation period (N=1389) | |
|---|---|---|---|---|
| | n (%) | Median Grams (Q1-Q3) | n (%) | Median Grams (Q1-Q3) |
| Amikacin | 6 (1.2%) | 480 (405-667.5) | 32 (2.3%) | 405 (117-705) |
| Amoxicillin | 8 (1.6%) | 2205 (1102.5-6990) | 30 (2.2%) | 1650 (990-4425) |
| Amoxicillin + clavulanic acid | 12 (2.3%) | 1575 (1100-4612.5) | 41 (3.0%) | 960 (480-1800) |
| Ampicillin | 22 (4.3%) | 2800 (1665-6825) | 104 (7.5%) | 4360 (2515-7020) |
| Ampicillin + Sulbactam | 35 (6.8%) | 6400 (4150-17725) | 191 (13.8%) | 6000 (3300-9600) |
| Azithromycin | 6 (1.2%) | 179 (150-839.5) | 22 (1.6%) | 204 (120-405) |
| Benzylpenicillin* | 12 (2.3%) | 4324000 (2160000-10680000) | 9 (0.6%) | 21000000 (18000000-24080000) |
| Benzathine benzylpenicillin* | 7 (1.4%) | 12000000 (8800000-19900000) | 2 (0.1%) | 4320000 (2880000-5760000) |
| Cephalexin | 14 (2.7%) | 1800 (1000-3850.5) | 11 (0.8%) | 800 (570-1750) |
| Cefotaxime | 2 (0.4%) | 258.5 (207.75-309.25) | 12 (0.9%) | 2440 (2070-4427.5) |
| Ceftazidime | 4 (0.8%) | 3450 (1575-5175) | 22 (1.6%) | 5130 (2250-7702.5) |
| Ceftazidime + avibactam | 2 (0.4%) | 13157 (10778.5-15535.5) | 6 (0.4%) | 6600 (3870-11130) |
| Ceftriaxone | 86 (16.7%) | 4000 (2000-7200) | 157 (11.3%) | 3920 (1920-7000) |
| Cefuroxime | 5 (1.0%) | 3600 (3000-11880) | 2 (0.1%) | 6180 (4770-7590) |
| Ciprofloxacin | 1 (0.2%) | 6000 (-) | 4 (0.3%) | 490 (307.5-825) |
| Clarithromycin | 78 (15.2%) | 1200 (720-2775) | 290 (20.9%) | 720 (400-1200) |
| Clindamycin | 38 (7.4%) | 2105 (1150.5-2962.5) | 42 (3.0%) | 2850 (1110-5370) |
| Gentamicin | 9 (1.8%) | 36 (15-65) | 70 (5.0%) | 78 (48-120) |
| Linezolid | 1 (0.2%) | 4320 (-) | 6 (0.4%) | 1560 (585-2463) |
| Meropenem | 28 (5.4%) | 1435 (666-3285) | 39 (2.8%) | 1950 (666-6030) |
| Metronidazole | 3 (0.6%) | 600 (321-1050) | 1 (0.1%) | 2400 (-) |
| Others | 13 (2.5%) | 2415 (1050-3600) | 28 (2.0%) | 3520 (945-11992.5) |
| Sodium Penicillin G* | 12 (2.3%) | 23000000 (8400000-40000000) | 52 (3.7%) | 11180000 (6450000-26600000) |
| Piperacillin + tazobactam | 51 (9.9%) | 10800 (4042.5-29335) | 91 (6.6%) | 10500 (4971-18000) |
| Trimethoprim + sulfamethoxazole | 15 (2.9%) | 960 (666-2016) | 43 (3.1%) | 320 (200-682) |
| Vancomycin | 40 (7.8%) | 1640 (600-3855) | 78 (5.6%) | 1730 (722.25-3810) |

*eTable 2. Healthcare-associated infections caused by antibiotic-resistant microorganisms during the study period.*

| Healthcare-associated infections | Baseline period n/N (Rate per 1,000) | Implementation period n/N (Rate per 1,000) | Ratio of Rates (CI 95%) | P-value |
|---|---|---|---|---|
| Infections caused by antibiotic-resistant microorganisms associated with urinary catheters | 0/603 (0.0) | 2/3270 (0.6) | - | - |
| Infections caused by antibiotic-resistant microorganisms associated with venous catheters | 0/962 (0.0) | 2/3433 (0.6) | - | - |
| Infections caused by antibiotic-resistant microorganisms associated with mechanical ventilation | 3/858 (3.5) | 2/2961 (0.7) | 0.19 (0.03;1.16) | 0.072 |
